# Serologic responses to SARS-CoV-2 infection among hospital staff with mild disease in eastern France

**DOI:** 10.1101/2020.05.19.20101832

**Authors:** Samira Fafi-Kremer, Timothée Bruel, Yoann Madec, Rebecca Grant, Laura Tondeur, Ludivine Grzelak, Isabelle Staropoli, François Anna, Philippe Souque, Catherine Schmidt-Mutter, Nicolas Collongues, Alexandre Bolle, Aurélie Velay, Nicolas Lefebvre, Marie Mielcarek, Nicolas Meyer, David Rey, Pierre Charneau, Bruno Hoen, Jérôme De Seze, Olivier Schwartz, Arnaud Fontanet

## Abstract

**Background:** The serologic response of individuals with mild forms of SARS-CoV-2 infection is poorly characterized.

**Methods:** Hospital staff who had recovered from mild forms of PCR-confirmed SARS-CoV-2 infection were tested for anti-SARS-CoV-2 antibodies using two assays: a rapid immunodiagnostic test (99.4% specificity) and the S-Flow assay (∼99% specificity).The neutralizing activity of the sera was tested with a pseudovirus-based assay.

**Results:** Of 162 hospital staff who participated in the investigation, 160 reported SARS-CoV-2 infection that had not required hospital admission and were included in these analyses. The median time from symptom onset to blood sample collection was 24 days (IQR: 21–28, range 13–39). The rapid immunodiagnostic test detected antibodies in 153 (95.6%) of the samples and the S-Flow assay in 159 (99.4%), failing to detect antibodies in one sample collected 18 days after symptom onset (the rapid test did not detect antibodies in that patient). Neutralizing antibodies (NAbs) were detected in 79%, 92% and 98% of samples collected 13–20, 21–27 and 28–41 days after symptom onset, respectively (P=0.02).

**Conclusion:** Antibodies against SARS-CoV-2 were detected in virtually all hospital staff sampled from 13 days after the onset of COVID-19 symptoms. This finding supports the use of serologic testing for the diagnosis of individuals who have recovered from SARS-CoV-2 infection. The neutralizing activity of the antibodies increased overtime. Future studies will help assess the persistence of the humoral response and its associated neutralization capacity in recovered patients.

## Introduction

A novel human coronavirus that is now named severe acute respiratory syndrome coronavirus 2 (SARS-CoV-2) emerged in Wuhan, China, in late 2019. In response, many countries have implemented large scale public health and social measures in an attempt to reduce transmission and minimize the impact of the outbreak. As the benefits of these measures are now becoming apparent in terms of a reduction in the daily incidence of SARS-CoV-2 infections and associated deaths, countries are looking for ways to lift these measures and resume economic and social activities. Ideally, the lifting of measures would occur if the population had built sufficient collective immunity, known as herd immunity, to the point that any reintroduction of the virus would not trigger a new epidemic wave. In this context, it is important to understand the extent to which infection has spread in communities, and to which those who have been infected may be protected from re-infection. This requires further understanding of antibody kinetics following SARS-CoV-2 infection.

Numerous serologic assays are now available, which provide information on extent of infection and estimates of protective immunity – that is, protection against re-infection. To date, it is thought that for hospitalised patients with COVID-19, seroconversion occurs within the second week following onset of symptoms, with a median time of 5–12 days for IgM antibodies and 14 days for IgG and IgA [1–6]. However, it remains unclear whether time to seroconversion may differ according to disease severity, and early reports suggest that individuals with mild infection may have delayed or absent seroconversion [4]. Further, the correlation between detection of antibodies generated in response to SARS-CoV-2 infection and protective immunity has not yet been established.

The first three COVID-19 cases identified in France were reported on 24 January 2020 in travellers returning from Wuhan, China [7]. Between 17 and 24 February, an annual religious gathering attended by 2500 people took place in Mulhouse, eastern France and resulted in a SARS-CoV-2 superspreading event. Infected individuals went to regional hospitals, and this led to a cluster of infected staff at the Strasbourg University Hospitals from the first week of March. Most of them are young individuals who developed mild forms of disease.

The epidemic in Strasbourg, and specifically, the cluster of infected hospital staff, provides the opportunity to use serologic assays, to assess antibody kinetics in individuals who had recovered from COVID-19 and to understand how this correlates with protective immunity.

## Methods

### Participants

Between 6 April and 8 April 2020, all hospital staff from Strasbourg University Hospitals with PCR-confirmed SARS-CoV-2 infection were invited to participate in the investigation. This invitation included doctors, nurses, physiotherapists, dentists, medical students, orderlies, hospital assistants, and hospital administrative staff.

Following informed consent, participants completed a questionnaire which covered sociodemographic information, underlying medical conditions, and details related to SARSCoV-2 infection, including date of testing, date of symptom onset and a description of symptoms. A 5 mL blood sample was taken from all participants.

### Serologic response measurement

All serum samples were tested for antibody responses to SARS-CoV-2 using two serologic assays: 1) a rapid immunodiagnostic assay detecting SARS-CoV-2 spike protein receptor binding domain (RBD) developed by Biosynex®; 2) the S-Flow assay, a flow-cytometry based assay that measures antibodies binding to the Spike protein (S) expressed at the surface of target cells [8]. Two parameters can be calculated with this assay: the first is the percentage of cells having captured antibodies, defining the seropositivity. The second is the mean fluorescence intensity (MFI) of this binding, which provides a quantitative measurement of the amount of antibodies and their efficacy [8]. As a control for the S-Flow tests, we included samples from pre-epidemics individuals, providing cut-offs for the the S-Flow >99% specificity ([8] and Fig. 1A). The rapid immunodiagnostic assay has a specificity estimated at 99.4% for the IgM, 100% for the IgG, and 99.4% for the combined IgM/IgG results (S.F.K., personal communication). Samples were also tested for neutralization activity using a viral pseudotype-based assay [8]. Briefly, single cycle lentiviral pseudotypes coated with the S protein and encoding for a luciferase reporter gene were preincubated with the serum to be tested at a dilution of 1:100, and added to 293T-ACE2 target cells. The luciferase signal was measured after 48h. The percentage of neutralization was calculated by comparing the signal obtained with each serum to the signal generated by control negative sera. In some analyses, we categorized the samples according to the extent of neutralization observed at the 1:100 dilution. Neutralizing activities >50% and >80% corresponded to inhibitory dilution 50% (ID50) >100 and ID80 >100, respectively.

**Figure 1.**
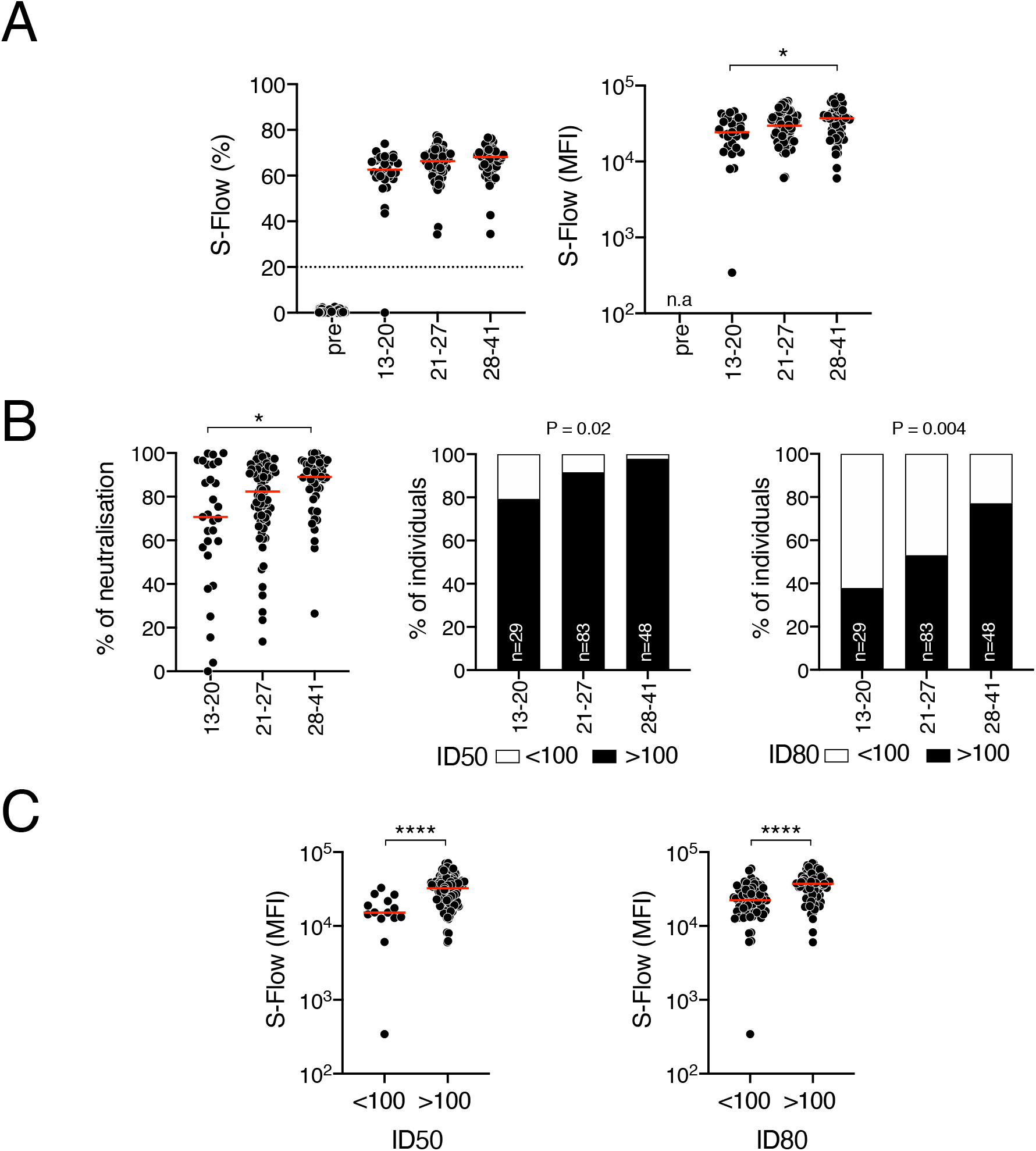
Analysis of SARS-Cov-2 antibody response. **(A)** Sera from the 160 HCW were surveyed for anti-SARS-Cov-2 antibodies. S-Flow data are represented by the frequency of S+ cells (n=160, left panel) and the median Fluorescence intensity (MFI) in positive samples (n=159, middle panel). Historical pre-epidemic samples (pre) were included to determine backgrounds of S Flow (n=140). Each dot represents a sample. Samples were grouped according to the number of days after symptom onset. Statistical analyses were performed using Kruskall-Wallis with Dunn’s multiple comparisons test or chi-2 test. * p<0.005. n.a.: not applicable **(B)** Neutralizing activity of the 160 sera. The ability of each serum to neutralize lentiviral S-pseudotypes was assessed at a serum dilution of 1:100 (left panel). Samples are grouped according to the number of days after symptom onset. For each time group, the frequencies of samples displaying a ID50 > 100 (middle panel) or a ID80 > 100 (left panel) were determined. Each dot represents a sample. ** p<0.005 Kruskall-Wallis with Dunn’s multiple comparisons test. **(C)** Relationship between serological measurement and neutralizing activity. The S-Flow MFI of samples displaying ID50 and ID80 above or below 100 are depicted. Each dot represents a sample. **** p<0.0001 Unpaired t-test. The statistically significant differences are depicted.

**Figure 2.**
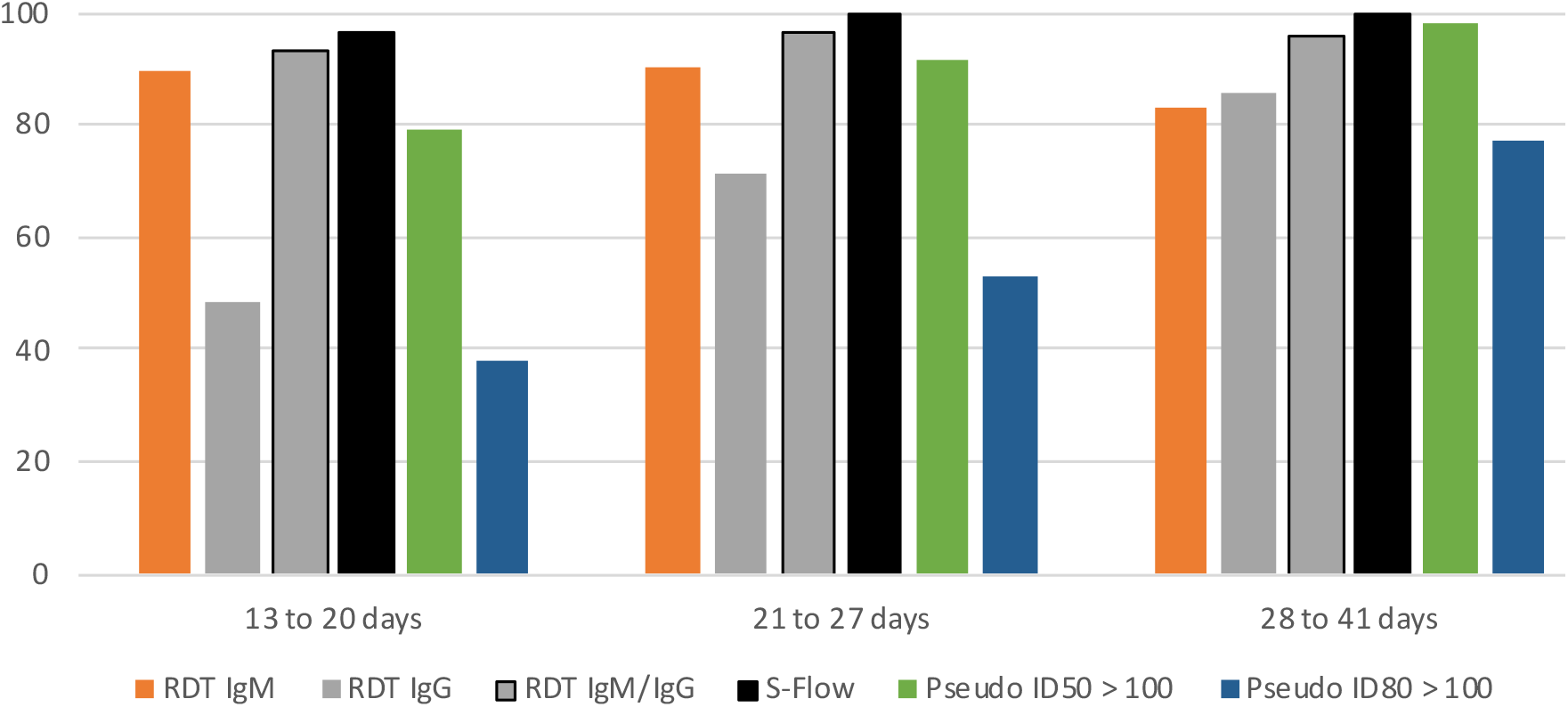
Seropositivity by serologic assay used (Rapid test, S-Flow, and pseudoneutralisation) according to the time between onset of symptoms and collection of blood sample.

### Statistical analyses

Seropositivity was defined as the presence of detectable anti-SARS-CoV-2 antibodies. The proportion of seropositive samples was compared by time between onset of symptoms and collection of blood sample using chi-square test.

Antibody neutralizing activity was compared by age, gender, underlying medical conditions, time from symptom onset and type of symptoms using chi-square or Fisher’s exact test where appropriate. Logistic regression was used for multivariable analysis.

The S-Flow MFI and neutralization of sera were compared by delay since onset of symptoms using the Kruskall-Wallis non-parametric test. The S-Flow MFI of sera with ID50 and ID80 above or below 100 were compared using Student’s t-test. The chi-2 test was used to evaluate the association between investigated factors and neutralization levels.

All analyses were performed using Stata (Stata Corp., College Station, Texas, USA) or GraphPad Prism 8 (GraphPad Software, LLC).

### Ethical considerations

This study was registered with ClinicalTrials.gov (NCT04325646) and received ethical approval by the Comité de Protection des Personnes Ile de France III. Informed consent was obtained from all participants.

## Results

Between 6 April and 8 April 2020, 162 hospital staff from Strasbourg University Hospitals who had recovered from RT-PCR confirmed SARS-CoV-2 infection participated in the investigation. Two individuals who were hospitalized for COVID-19 were excluded from these analyses to determine serologic responses in those with mild forms of COVID-19. Table 1 indicates the characteristics of these 160 hospital staff. The median age was 32 years (inter quartile range (IQR): 26–44) and 50 (31.2%) were males. The majority of participants were medical students (28.1%), doctors (20.0%) or nurses (19.4%).

**Table 1.**
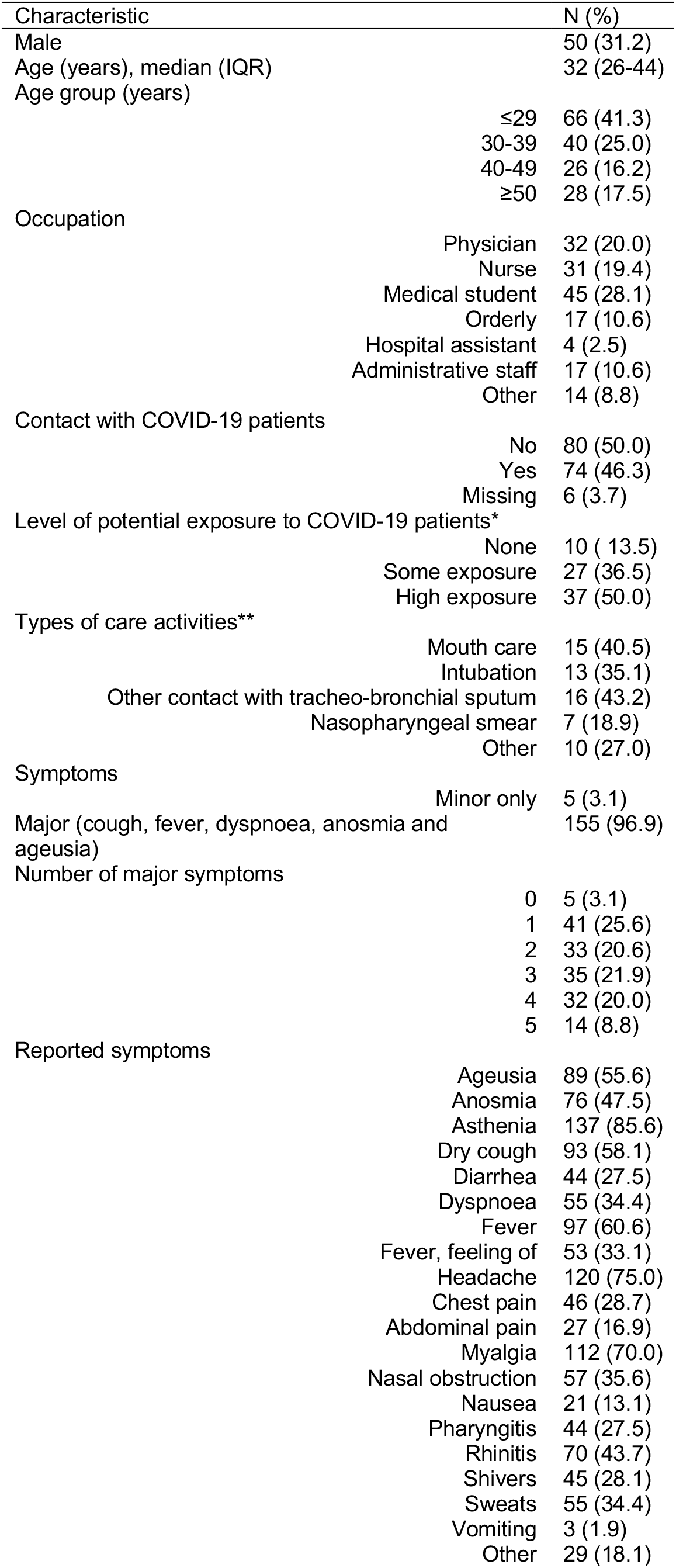
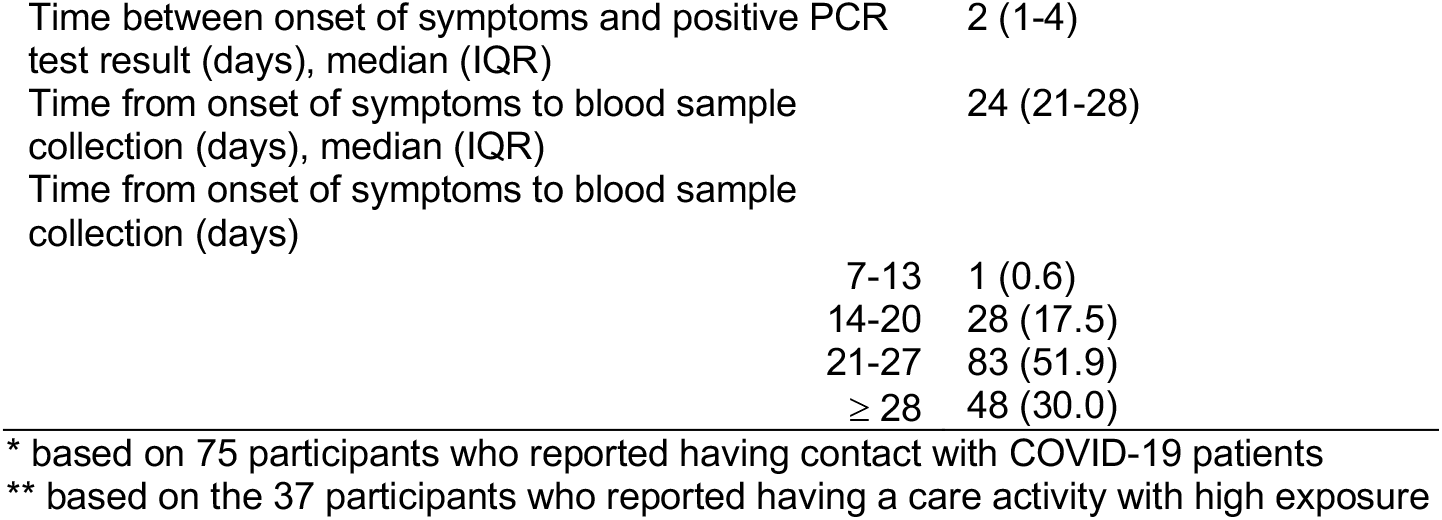
Characteristics of the 160 hospital staff with PCR-confirmed SARS-CoV-2 infection

In terms of possible sources of SARS-CoV-2 infection, 74 (46.2%) reported having had contact with a COVID-19 patient either in the ward or in the emergency room. A further 38 (23.7%) reported having had contact with a COVID-19 case outside the health care setting.

One hundred and fifty five (96.9%) had symptoms consistent with COVID-19 (dry cough, fever, dyspnea, anosmia or ageusia). The median time between onset of symptoms and PCR testing was 2 days (IQR:1–4), and the median time from onset of symptoms to blood sampling was 24 days (IQR: 21–28, range 13–39).

Figure 1 and Table 2 indicate the seropositivity rates detected by the three assays and categorized by the delay between onset of symptoms and collection of samples. Across all 160 participants, 159 had detectable anti-SARS-CoV-2 antibodies by S-Flow (99.4% sensitivity). The only participant whose serology was negative with all assays was a 58-year-old male with a body mass index of 32 kg/m^2^ and no other risk factors for severe COVID-19 disease. His blood was sampled 18 days after onset of symptoms which persisted at the time of blood collection. The S-Flow MFI displays a significantly higher signal in individuals sampled at days 28–41 compared to those sampled at days 13–20 (Figure 1A). These results suggest that the overall amount or the affinity of the antibodies improved with time since onset of symptoms.

**Table 2.** Seropositivity with the different assays (Rapid test, S-Flow, and pseudoneutralisation) according to the time after onset of symptoms

| Time from onset of symptoms (days) | 13-20<br>(n=29) | 21-27<br>(n=83) | ≥28<br>(n=48) | Total | P value |
| --- | --- | --- | --- | --- | --- |
| Rapid test IgM | 26 (89.7) | 75 (90.4) | 40 (83.3) | 141 (88.1) | 0.47 |
| Rapid test IgG | 14 (48.3) | 59 (71.1) | 41 (85.4) | 114 (71.2) | 0.002 |
| Rapid test IgG or IgM | 27 (93.1) | 80 (96.4) | 46 (95.8) | 153 (95.6) | 0.76 |
| S-Flow | 28 (96.5) | 83 (100) | 48 (100) | 159 (99.4) | 0.18 |
| Pseudoneutralisation ID50 >100 | 23 (79.3) | 76 (91.6) | 47 (97.9) | 146 (91.2) | 0.020 |
| Pseudoneutralisation ID80 >100 | 11 (37.9) | 44 (53.0) | 37 (77.1) | 92 (57.5) | 0.002 |

The IgM rapid test appeared more sensitive than IgG (overall sensitivity: 88.1% vs 71.2%, repectively), especially at the earlier timepoints (Table 2). The combination of IgG and IgM rapid test data increased the sensitivity to 95.6%.

Figure 1B and Table 3 show the proportion of individuals with a neutralizing activity, using the pseudovirus neutralization assay. The proprotion of samples with neutralizing activity increased over time (Figure 1B), reflecting the increase of antibody titers observed with the SFlow. The proportion of individuals with an ID50 ≥100 were 79 %, 92% and 98% at 13–20, 21–27 and 28–41 days after symptom onset, respectively (P=0.02) (Figure 1B).

**Table 3.** Proportion of 160 participants with protective immunity according to time since onset of symptoms, type of symptoms, age, underlying medical conditions and tobacco use.

|  |  | N | Neutralization<br>ID50 > 100 | P value | Neutralization<br>ID80 > 100 | P value |
| --- | --- | --- | --- | --- | --- | --- |
| Time between onset of symptoms and collection of blood sample (days) |  |  |  | 0.02 |  | 0.004 |
|  | 13-20 | 29 | 23 (79.3) |  | 11 (37.9) |  |
|  | 21-27 | 83 | 76 (91.6) |  | 44 (53.0) |  |
|  | ≥28 | 48 | 47 (97.9) |  | 37 (77.0) |  |
| Number of participants with major symptoms |  |  |  | 0.87 |  | 0.44 |
|  | 0 | 5 | 4 (80.0) |  | 3 (60.0) |  |
|  | 1 | 41 | 38 (92.7) |  | 29 (70.7) |  |
|  | 2 | 33 | 30 (90.9) |  | 16 (48.5) |  |
|  | 3 | 35 | 35 (94.3) |  | 20 (57.1) |  |
|  | 4 | 32 | 33 (94.3) |  | 16 (50.0) |  |
|  | 5 | 14 | 12 (85.7) |  | 8 (57.1) |  |
| Ageusia |  |  |  | 0.85 |  | 0.39 |
|  | No | 84 | 77 (91.7) |  | 51 (60.7) |  |
|  | Yes | 76 | 69 (90.8) |  | 41 (53.9) |  |
| Anosmia |  |  |  | 0.21 |  | 0.48 |
|  | No | 71 | 67 (94.4) |  | 43 (60.6) |  |
|  | Yes | 89 | 79 (88.8) |  | 49 (55.1) |  |
| Dry cough |  |  |  | 0.22 |  | 0.04 |
|  | No | 67 | 59 (88.1) |  | 45 (67.2) |  |
|  | Yes | 93 | 87 (93.5) |  | 47 (50.5) |  |
| Fever |  |  |  | 0.15 |  | 0.29 |
|  | No | 63 | 55 (87.3) |  | 33 (52.4) |  |
|  | Yes | 97 | 91 (93.8) |  | 59 (60.8) |  |
| Gender |  |  |  | 0.41 |  | 0.07 |
|  | Male | 50 | 47 (94.0) |  | 34 (68.0) |  |
|  | Female | 110 | 99 (90.0) |  | 58 (52.7) |  |
| Age group |  |  |  | 0.92 |  | 0.17 |
|  | ≤29 | 66 | 59 (89.4) |  | 33 (50.0) |  |
|  | 30-39 | 40 | 37 (92.5) |  | 23 (57.5) |  |
|  | 40-49 | 26 | 24 (92.3) |  | 15 (57.7) |  |
|  | ≥50 | 28 | 26 (92.9) |  | 21 (75.0) |  |
| BMI |  |  |  | 0.22 |  | 0.02 |
|  | <18.5 | 10 | 7 (70.0) |  | 3 (30.0) |  |
|  | 18.5-25 | 105 | 97 (92.4) |  | 55 (52.4) |  |
|  | 25-30 | 27 | 25 (92.6) |  | 19 (70.4) |  |
|  | ≥30 | 17 | 16 (94.1) |  | 14 (82.4) |  |
|  | Missing | 1 | 1 (100) |  | 1 (100) |  |
| Arterial hypertension |  |  |  | 0.31 |  | 0.03 |
|  | No | 150 | 136 (90.7) |  | 83 (55.3) |  |
|  | Yes | 10 | 10 (100) |  | 9 (90.0) |  |
| Asthma |  |  |  | 0.02 |  | 0.67 |
|  | No | 149 | 138 (92.6) |  | 85 (57.1) |  |
|  | Yes | 11 | 8 (72.7) |  | 7 (63.6) |  |
| Flu vaccine |  |  |  | 0.02 |  | 0.46 |
|  | No | 104 | 99 (95.2) |  | 62 (59.6) |  |
|  | Yes | 56 | 47 (83.9) |  | 30 (53.6) |  |
| Blood group |  |  |  | 0.26 |  | 0.96 |
|  | A | 55 | 50 (90.9) |  | 31 (56.4) |  |
|  | B | 18 | 18 (100) |  | 9 (50) |  |
|  | AB | 3 | 2 (66.7) |  | 2 (66.7) |  |
|  | O | 50 | 44 (88.0) |  | 30 (60.0) |  |
|  | Not specified | 34 | 32 (94.1) |  | 20 (58.8) |  |
| Tobacco use |  |  |  | 0.57 |  | 0.97 |
|  | No | 141 | 128 (90.8) |  | 81 (57.5) |  |
|  | Yes | 19 | 18 (94.7) |  | 11 (57.9) |  |
| Exposure to patients |  |  |  | 0.32 |  | 0.49 |
|  | None | 96 | 85 (88.5) |  | 52 (54.2) |  |
|  | Low | 27 | 26 (96.3) |  | 18 (66.7) |  |
|  | High | 37 | 35 (94.6) |  | 22 (59.5) |  |

The associations between the neutralizing activity and the type of symptoms, age, underlying medical conditions and tobacco use are summarized in Table 3. The characteristics associated with neutralizing activity (ID50 > 100) were time since onset of symptoms (P=0.02), absence of asthma (P=0.02), and absence of a flu vaccine (P=0.02). In a multivariable model including the three variables, none remained associated with neutralizing activity. We also analysed the association of high neutralizing activity (ID80≥100) with patients characteristics. High neutralizing activity was associated with time since onset of symptoms (P = 0.004), having a dry cough (P = 0.04), male gender (P=0.07), high BMI (P=0.02), and high blood pressure (P = 0.03). All these characteristics remained independently associated with high neutralizing activity in multivariable analysis except for high blood pressure (P = 0.11). There was no association between neutralizing activity and ageusia, anosmia, or fever.

We next examined the relationship between the extent of antibody response and the neutralizing capacity of the sera. Regardless of the time post-symptom onset, samples with ID50 and ID80 ≥100 displayed significantly higher signals in the S-Flow assay (Figure 1C).

## Discussion

In this investigation, we described the serologic responses of 160 hospital staff who recovered from PCR-confirmed mild SARS-CoV-2 infection. Most studies published to date have been based on hospitalized patients, and therefore have not been able to evaluate serologic responses in individuals with mild or subclinical infection. Since these individuals are currently understood to represent at least 80% of all SARS-CoV-2 infections [9], it is crucial to assess antibody responses in those with mild disease. In our study, we were able to show that all but one (99.4%) participant had detectable levels of anti-SARS-CoV-2 antibodies from 13 days after onset of symptoms. The differences observed between time to seroconversion across the different assays reflect their sensitivity. The S-Flow assay, which displays a high sensitivity, detected seroconversion in all but one sample. The rapid immunodiagnostic test performed well 21 days after onset of symptoms. The rapid test therefore has utility as a tool for diagnosis in the recovery phase of infection. The neutralization assay was positive in 91% of the samples, and the extent of neutralization paralleled the levels of signal obtained with the S-Flow and.

At the community level, countries that have implemented public health and social measures to limit transmission are now lifting some of these measures. Most of the evidence to date suggests that herd immunity after the first wave of the epidemic will be far from sufficient to provide protection against a second epidemic wave [10]. In our study, neutralizing ID50 ≥100 were found in 91% of the individuals. We further report that the neutralization activity of the serum increases with time, reaching 97% four weeks after the onset of symptoms. Therefore, it is a fair assumption that the majority of individuals with mild COVID-19 generate neutralizing antibodies within a month after onset of symptoms. Although not yet demonstrated, several lines of evidence suggest that the presence of neutralizing antibodies may be associated with protective immunity for SARS-CoV-2 infection. In humans, passive immunotherapy based on transfer of antibodies from recovered COVID-19 patients decreases disease severity [1,3,6]. In a monkey model, protection from a second SARS-CoV-2 infection is associated with the presence of neutralizing antibodies in the serum [11]. SARS-CoV-2 NAbs are known to be present in symptomatic individuals [5,12–14]. In a study of 175 convalescent patients with mild symptoms, NAbs were most often detected 10–15 days after symptom onset [14]. However, about 30% of recovered patients generated low titers of NAbs (≤1:500), even at a later time point [14]. Our results are in line with this observation and indicate that recovery from mild cases is generally, but not always, associated with high titers of NAbs in the serum. Indeed, we report here that one month after the onset of symptoms, 98% and 77% of individuals display Nabs with an ID50 and ID80 ≥100, repectively. Antibody titers are generally higher in patients with severe or critical diseases [6,14]. Interestingly, in our study, individuals with factors associated with more severe disease (e.g., male sex, high body mass index and high blood pressure), were more likely to have high titers of neutralizing antibodies compared to others. This may be due to a higher antigenic burden in such individuals, which will generate a stronger humoral response, or may, on the contrary, suggest that some antibodies may play a deleterious role during infection [15]. Future studies are warranted to characterize the beneficial or detrimental role of specific antibodies in COVID-19 patients and the minimal titer required for protection.

For patients with SARS-CoV-1, antibodies persist for at least 2 years after symptomatic infection [16]. In the case of Middle East Respiratory Syndrome (MERS)-CoV, the antibody response is variable, not robust, and often undetectable when disease is mild [17–20]. Future studies will help evaluating the persistence of antibodies upon SARS-CoV-2 infection. The cohort of hospital staff described here provides the opportunity to study the duration of the humoral response and the dynamics of the neutralization capacity of the sera. A clinical and virological assessment of potential reinfections will also help establishing the links that may exist between the antibody response and immune protection.

## Data Availability

All data generated or analysed during this study are included in this published article (and its supplementary information files).

## Acknowledgments

We thank the patients and individuals who donated their blood and the ICAReB team for management and distribution of the samples.

## Authors contribution

Conceptualization and Methodology: SFK, TB, YM, OS, AF

Cohort management and sample collection: SFK, YM, RG, LT, CSM, NC, AB, AV, NL, MM, NM, DR, BH, JDS, AF

Serological and seroneutralisation assays: TB, LG, IS, FA, PS, SVDW, PC, OS

Data assembly and manuscript writing: SFK, TB, YM, RG, LT, OS, AF

Funding acquisition: PC, TR, BH, JDS, OS, AF

Supervision: OS, AF

## Conflicts of interest

SFK, TB, YM, RG, LT, LG, IS, FA, PS, CSM, NC, AB, AV, NL, MM, NM, DR, BH, JDS, OS and AF have no competing interest to declare.

PC is the founder and CSO of TheraVectys.

## Fundings

OS lab is funded by Institut Pasteur, ANRS, Sidaction, the Vaccine Research Institute (ANR-10-LABX-77), Labex IBEID (ANR-10-LABX-62-IBEID), “TIMTAMDEN” ANR-14-CE14–0029, “CHIKV-Viro-Immuno” ANR-14-CE14–0015–01 and the Gilead HIV cure program. LG is supported by the French Ministry of Higher Education, Research and Innovation. SFK lab receives funding from Strasbourg University Hospitals (COVID-HUS; CE-2020–34)

## Notes

### Clinical Trial

NCT04325646

### Author Declarations

This study was registered with ClinicalTrials.gov (NCT04325646) and received ethical approval by the Comite de Protection des Personnes Ile de France III. Informed consent was obtained from all participants.

